# Endemic Chikungunya Fever in Kenyan Children

**DOI:** 10.1101/2020.07.22.20116707

**Authors:** Doris K. Nyamwaya, Mark Otiende, Donwilliams O. Omuoyo, George Githinji, Henry K. Karanja, John N. Gitonga, Zaydah de Laurent, James R. Otieno, Rosemary Sang, Everlyn Kamau, Stanley Cheruiyot, Edward Otieno, Charles N. Agoti, Philip Bejon, Samuel M. Thumbi, George M. Warimwe

**Author notes:** Correspondence: George Warimwe, KEMRI-Wellcome Trust Research Programme, P.O. Box 230-80108, Kilifi, Kenya. contributed equally (Doris K. Nyamwaya, Mark Otiende).

## Abstract

**Background:** Chikungunya virus (CHIKV) was first identified in Tanzania in 1952. Several epidemics including East Africa are described, but there are no descriptions of longitudinal surveillance of endemic disease. Here, we estimate the incidence of CHIKV and describe viral phylogeny in coastal Kenya.

**Methods:** Over a 5-year period (2014-2018), 11,708 febrile illnesses in 5,569 children visiting two primary healthcare facilities linked to a demographic surveillance system in coastal Kenya were recorded and blood samples obtained. Reverse-transcriptase PCR was used to identify CHIKF cases in 3,500 children randomly selected from the 5,569 children.

**Results:** We found CHIKF to be endemic in this setting, associated with 12.7% (95% CI 11.60, 13.80) of all febrile presentations to primary healthcare. The prevalence of CHIKV infections among asymptomatic children in a community survey was 0.7% (95% CI 0.22, 2.12). CHIKF incidence among children <1 year of age was 1703 cases/100,000-person years and 46 cases/100,000-person years among children aged ≥ 10 years. Recurrent CHIKF episodes, associated with fever and viraemia, were observed among 19 of 170 children with multiple febrile episodes during the study period and confirmed by genome sequencing. All sequenced viral genomes mapped to the ECSA genotype albeit distinct from CHIKV strains associated with the 2004 East African epidemic.

**Conclusions:** CHIKF may be a substantial public health burden in primary healthcare on the East African coast outside epidemic years, and recurrent infections are common.

## BACKGROUND

Chikungunya fever (CHIKF) is a zoonotic mosquito-borne febrile illness characterised by acute, often chronic, debilitating polyarthralgia and polyarthritis that can last for months to years [1-3]. The disease is caused by chikungunya virus (CHIKV), a positive sense RNA virus in the family *Togaviridae* that was first isolated from a febrile patient in Tanzania in 1952 [4]. CHIKV transmission between humans is mainly mediated by the geographically widespread *Aedes aegypti* and *Ae. albopictus* mosquitoes, with geographic spread and spillover from sylvatic transmission cycles in monkeys thought to account for the periodic re-emergence of human disease outbreaks [5-8].

The CHIKV genome is approximately 11.8kb in length and encodes four non-structural proteins (nsP1 to nsP4) required for viral replication, three major structural proteins (the capsid protein, and envelope proteins E1 and E2), and two small polypeptides (6K/TF and E3) [9]. Exposure to CHIKV results in acquisition of protective virus neutralising antibodies that target the E2 protein, and is the basis for ongoing efforts to develop CHIKF vaccines [10, 11]. Phylogenetic studies have defined three CHIKV genotypes, namely East, Central and Southern Africa (ECSA), West Africa, and the Asian genotype [12]. In 2004, CHIKV re-emerged in coastal Kenya causing one of the largest epidemics on record, affecting millions of people as it spread along the Indian Ocean islands, India, southeast Asia and Europe [8, 13, 14]. This epidemic was associated with emergence of the Indian Ocean Lineage (IOL), which mapped within the ECSA genotype and included viruses with adaptive mutations in the E1 protein that increased their transmissibility by *Aedes albopictus* mosquitoes [15, 16].

CHIKF epidemics are now frequently reported globally [8, 17], but despite its original discovery and later re-emergence in East Africa, very little is known regarding inter-epidemic CHIKV exposure in the region. CHIKF cases have previously been detected in children in Kenya and Tanzania during inter-epidemic periods [18-20], which together with the high anti-CHIKV antibody seroprevalence observed in these settings suggest endemic CHIKV transmission [21, 22]. To address these knowledge gaps, we conducted a primary healthcare-based study linked to demographic surveillance and community survey to estimate the prevalence and incidence of CHIKV infections among children in Kilifi, coastal Kenya. Our study period, 2014 to 2018, included the most recent CHIKF epidemic year, 2016 [23], allowing an unprecedented assessment of exposure risk and disease burden before, during and after a CHIKF epidemic in this setting.

## METHODS

### Study population

We conducted a population-based observational cohort study between March 2014 and October 2018 at two dispensaries, Ngerenya and Pingilikani, based within the Kilifi Health Demographic Surveillance System (KHDSS) [24]. The KHDSS covers an area of 891km^2^ with approximately 290,000 residents that are enumerated every four months at the household level [24]. Ngerenya and Pingilikani have different rates of mosquito-borne pathogen exposure as inferred from previous malaria studies; Ngerenya and environs experiences low intensity malaria transmission, whereas malaria transmission around Pingilikani is moderate [25]. However, no studies on arbovirus exposure had been conducted previously. Our approach was to screen children presenting with fever (axillary temperature of ≥37.5°C) at the dispensaries for CHIKV infection by reverse-transcriptase polymerase chain reaction (RT-PCR). Children presenting at the dispensaries were linked to the KHDSS data using a unique identifier, allowing us to estimate incidence in the population, and to determine the influence of various demographic variables on the incidence estimates. Ethical approval was provided by the Kenya Medical Research Institute Scientific and Ethics Review Unit (SSC Nos. 3296, 2617 and 3149).

### CHIKV RT-PCR assays

CHIKV infection was defined as detection of CHIKV viral RNA by RT-PCR using a published primer-probe set targeting the CHIKV non-structural protein 1 (nsP1) region[26], namely CHIKV 874 (5’-AAAGGGCAAACTCAGCTTCAC-3’), CHIKV 961 (5’-GCCTGGGCTCATCGTTATTC-3’) and CHIKV 899-FAM (5’-*FAM*-CGCTGTGATACAGTGGTTTCGTGTG-*TAMRA*-3’). Briefly, 100µl of finger-prick blood samples collected at presentation to the dispensaries, and stored at −80°C, was used for total RNA isolation using TRIzol™ Reagent (ThermoFisher) as per manufacturer instructions. Presence of CHIKV viral RNA was then determined using the QuantiFast RT-PCR Kit (Qiagen) in a 25µl reaction with primers and probe at a final concentration of 100nM and 20nM, respectively, and 5µl total RNA template. RT-PCR assays were done on a 7500 Real-Time PCR System (Applied Biosystems) with cycling conditions as follows: 50°C for 20 minutes, 95°C for 15 minutes, followed by 45 cycles of 94°C for 15 seconds and 60°C for 1 minute. A second confirmatory screen was performed on some of the RT-PCR positive samples using an assay targeting the CHIKV nsP4 region as previously described[27]. For both assays, a positive result was defined as a cycle threshold (Ct) value of <40. Viral RNA from a cultured CHIKV isolate obtained from a febrile Kenyan patient was used as a positive control, while RT-PCR mastermix without template was used as a negative control.

### CHIKV genome sequencing and analysis

RT-PCR positive CHIKV samples were sequenced using the Nanopore MinION technology following PCR amplification using methods described in the PrimalSeq approach^1 7^. Full details of our sequencing and bioinformatics workflow can be found in the Supplementary Material. In brief, CHIKV-specific multiplex primers were designed using the Primal Scheme software and used for pre-enrichment of CHIKV viral RNA in the samples using a multiplex PCR method. Barcodes and sequencing adapters were ligated into each sample, after which all samples were pooled into a single reaction tube and the library loaded on to a MinION sequencing device for sequencing. The sequences generated in this study (N=10) were deposited in GenBank, accession numbers: MT526796-MT526807. MUSCLE was used to align the Kilifi CHIKV genome sequences with those available in GenBank from other geographical locations (see Supplementary Material). Maximum Likelihood phylogenies were reconstructed from the alignment using RAxML[28] using the GTR substitution model with 4 gamma categories (GTR+G4) and visualized in FigTree v1.4.4.

### Sampling and statistical analyses

We selected a random sample of 3500 out of 5,669 children who visited the two dispensaries with febrile illness. Associations between available demographic variables and CHIKV RT-PCR positivity were assessed using odds ratios estimated using logistic regression models. CHIKF incidence within the dispensary catchment area, defined as radius of 5km from each dispensary, was expressed per 100,000 person-years observed and incidence rate ratios (IRRs) comparing various demographic variables determined by Poisson regression. Univariate analyses were performed using Chi2 test (categorical variables) or the Mann-Whitney U test (continuous variables). All statistical analyses were carried out in Stata™ version 15 with a two-sided p value <0.05 used for statistical significance.

## RESULTS

### CHIKV infections are common among children in coastal Kenya

Between March 2014 and October 2018, there were 29,819 visits by children aged up to 15 years to the two dispensaries (6,746 in Ngerenya, 23,073 in Pingilikani), of which 13,696 were febrile (Figure 1). Of the 13,696 fevers, 1,256 lacked a unique patient identifier and 732 samples were missing. After these exclusions, there were 11,708 febrile visits from a total of 5,569 children (median 1 febrile visit per child during the study period, IQR 1 - 2) eligible for CHIKV RT-PCR analysis (Figure 1).

**Figure 1:**
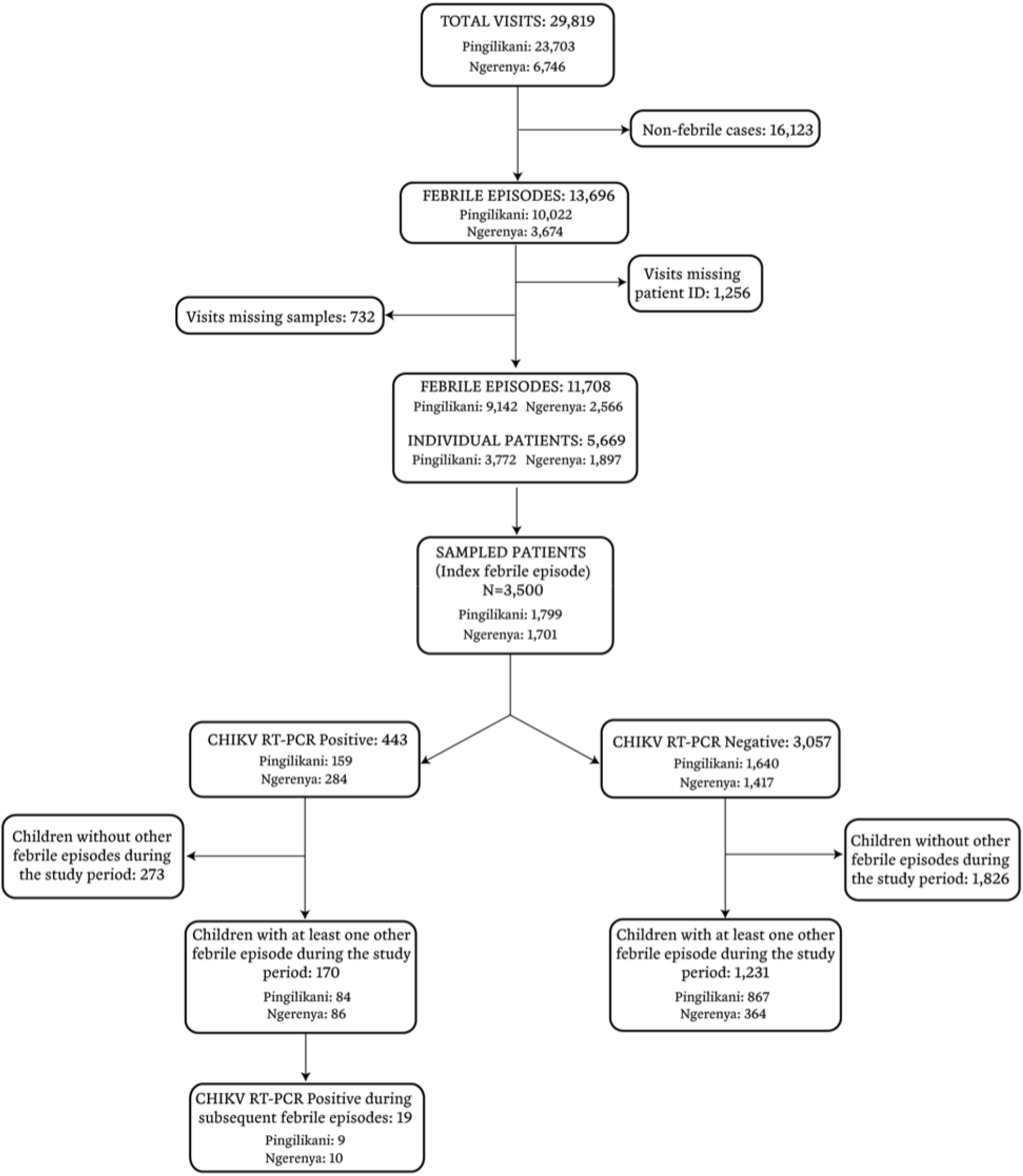
Study population. The flow of study participants, reasons for exclusion and results from CHIK RT-PCR screening are shown.

A sample of 3,500 febrile children out of the 5,569 across the two dispensaries were randomly selected and samples from their first (index) febrile episode during the study period screened for CHIKV infection (Figure 1). The median age of these 3,500 children was 3.1 years (IQR 1.3 – 6.4), with 1,701 being resident in Ngerenya and 1,799 in Pingilikani, respectively (Figure 1). Our study period, 2014 to 2018, was at least 10 years since the 2004 CHIKF epidemic occurred in Kenya, with ∼90% our study population having been born after that epidemic.

Of the 3,500 children 443 were RT-PCR positive (12.7%, 95% CI 11.60, 13.80; Table 1), with RT-PCR positivity being twice as frequent in Ngerenya (16.7%, 95% CI 15.00, 18.54) than in Pingilikani (8.8%, 95% CI 7.61, 10.24). Most of the 443 CHIKF cases had a clinical diagnosis of upper respiratory tract infection (URTI) but in no instance was a diagnosis of CHIKF assigned (Table 1). The treatments prescribed for the management of CHIKF cases were comparable to those for other causes of fever at the dispensaries, but fewer antimalarials were prescribed for CHIKF cases (Table 1). CHIKF case prevalence was highest in 2016 (23.1%), when an epidemic was reported in Kenya [23], and was significantly lower in the pre- and post-epidemic years where it ranged between 5% and 8% (Table 1). In contrast, the prevalence of asymptomatic CHIKV infections estimated by cross-sectional sampling of a different group of 435 children in the same study location in 2016 was 0.7% (95% CI 0.22, 2.12); this yielded a clinical-to-asymptomatic CHIKV infection ratio of 18:1 during the epidemic year.

**Table 1.**
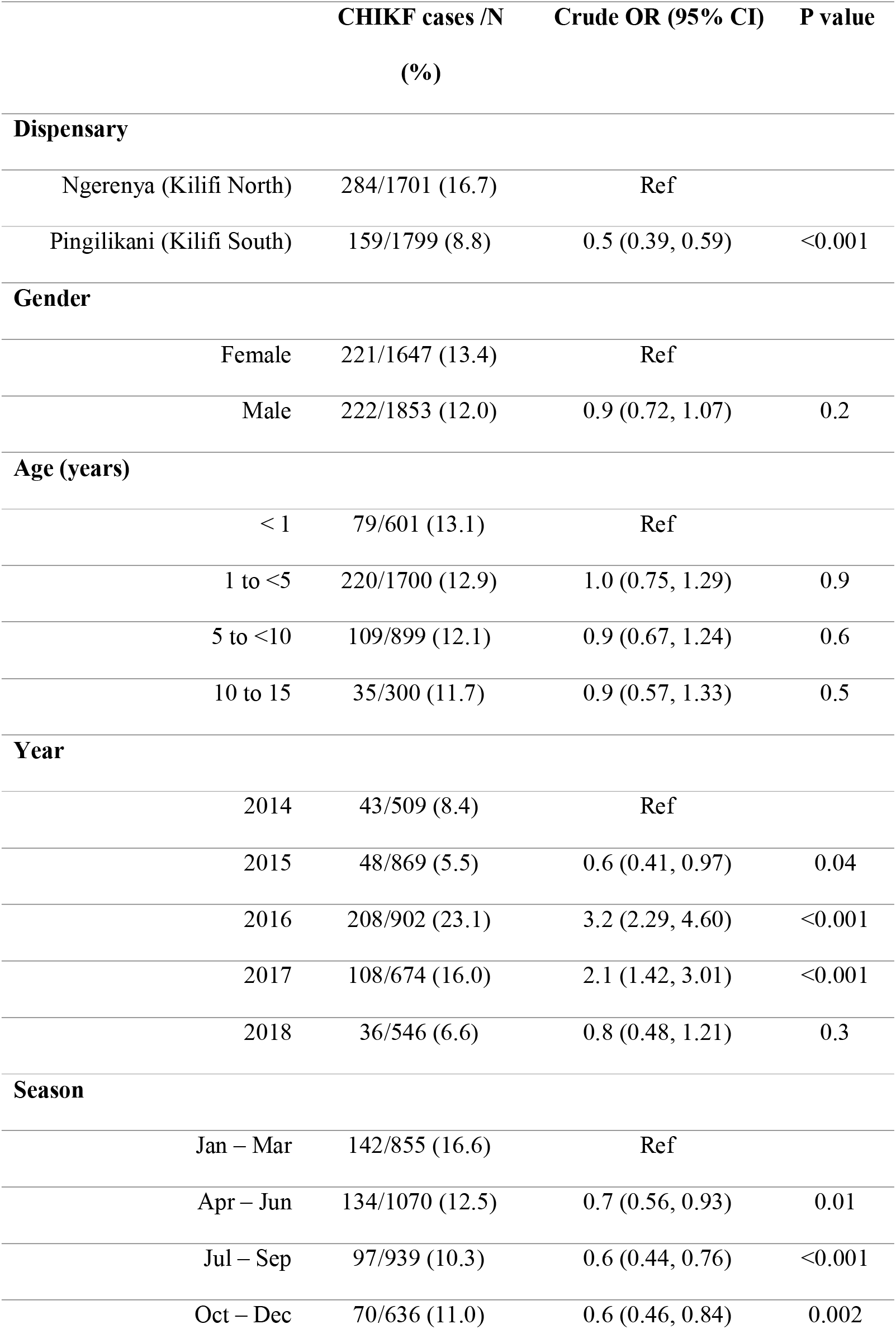
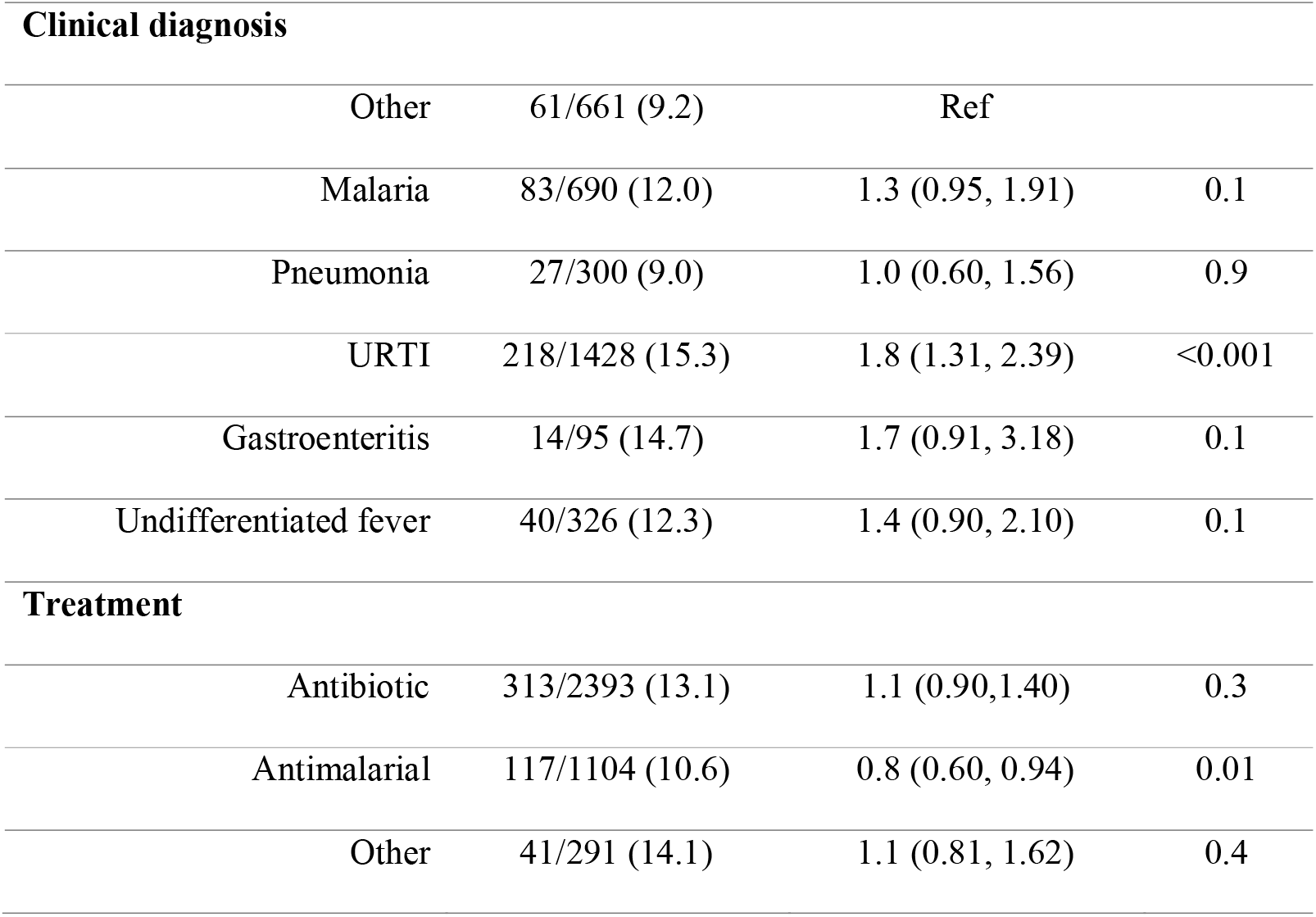
Demographic characteristics of study participants.

### Incidence of CHIKF in the community

We calculated the incidence of CHIKF restricting our analysis to children aged up to 15 years whose residence was within the dispensary catchment area. This represented a total of 139,840 person-years of observation (pyo) over the study period at both locations. For the numerator (CHIKF cases), we applied the observed CHIKV RT-PCR positivity rates (Table 1) to the total number of febrile children presenting to the respective dispensary on the assumption that the case positive rate among the 3,500 we randomly selected would hold true for the remaining 2,069 samples.

The incidence of CHIKF during the study period was 340 cases/100,000 pyo (95% CI 310, 373; Table 2). However, an inverse relationship was evident between CHIKF incidence and age, consistent with acquisition of protective immunity against disease due to ongoing CHIKV exposure in this setting (Table 2). CHIKF incidence was significantly higher in Ngerenya as compared to Pingilikani; furthermore, while an increase in incidence was observed in Ngerenya during the 2016 CHIKF epidemic, such an increase was not observed in Pingilikani where CHIKF incidence had been gradually declining since 2014 (Table 2). Marked seasonality in CHIKF incidence was observed in Pingilikani but not Ngerenya (Table 2) suggesting potential differences in ecological risk factors of disease. A negative correlation was observed between CHIKF incidence and residential distance from the dispensary at both locations, which likely reflected access to care (Table 2) as has been previously reported for malaria incidence [29].

**Table 2.** Incidence of CHIKF in the study population.

|  | **Ngerenya** |  | **Pingilikani** |  |
| --- | --- | --- | --- | --- |
|  | **N=74,799 person-years** |  | **N=65,041 person-years** |  |
|  | **Incidence** | **IRRs** | **Incidence** | **IRRs** |
|  | **(95% CI)** | **(95% CI)** | **(95% CI)** | **(95% CI)** |
| **Overall incidence** | 408 (363, 457) | - | 267 (228, 311) | - |
| **Year** |  |  |  |  |
| 2014 | - | - | 643 (513, 794) | Ref |
| 2015 | 67 (32, 123) | Ref | 289 (205, 397) | 0.43 (0.30, 0.64) |
| 2016 | 1205 (1035, 1395) | 18.08 (9.56, 34.18) | 187 (120, 279) | 0.28 (0.18, 0.44) |
| 2017 | 611 (493, 750) | 9.17 (4.77, 17.61) | 62 (27, 122) | 0.09 (0.05, 0.19) |
| 2018 | 104 (56, 178) | 1.57 (0.69, 3.57) | 93 (44, 170) | 0.14 (0.07, 0.27) |
| **Gender** |  |  |  |  |
| Female | 382 (321, 452) | Ref | 328 (268, 398) | Ref |
| Male | 432 (368, 505) | 1.13 (0.90, 1.42) | 207 (160, 264) | 0.63 (0.46, 0.86) |
| **Age (years)** |  |  |  |  |
| < 1 | 2209 (1776, 2715) | Ref | 1198 (856, 1631) | Ref |
| 1 to <5 | 791 (670, 927) | 0.36 (0.28, 0.46) | 587 (476, 717) | 0.49 (0.34, 0.71) |
| 5 to <10 | 198 (146, 263) | 0.09 (0.06, 0.13) | 76 (44, 124) | 0.06 (0.04, 0.11) |
| 10 to 15 | 24 (9, 52) | 0.01 (0.005, 0.02) | 68 (38, 112) | 0.06 (0.03, 0.10) |
| **Distance from**<br>**dispensary (Km)** |  |  |  |  |
| <1 | 800 (531, 1156) | Ref | 560 (306, 939) | Ref |
| 1 to <2 | 930 (742, 1152) | 1.16 (0.76, 1.79) | 1038 (760, 1384) | 1.85 (1.02, 3.37) |
| 2 to <3 | 657 (531, 804) | 0.82 (0.54, 1.25) | 837 (658, 1049) | 1.50 (0.85, 2.65) |
| 3 to <4 | 513 (396, 654) | 0.64 (0.41, 1.00) | 224 (128, 363) | 0.40 (0.20, 0.82) |
| 4 to <5 | 118 (70, 186) | 0.15 (0.08, 0.27) | 118 (59, 211) | 0.21 (0.10, 0.46) |
| 5 to <6 | 34 (13, 74) | 0.04 (0.02, 0.10) | 20 (7, 43) | 0.04 (0.01, 0.09) |
| **Season** |  |  |  |  |
| Jan - Mar | 463 (371, 572) | Ref | 227 (160, 312) | Ref |
| Apr - Jun | 348 (269, 444) | 0.75 (0.54, 1.04) | 528 (422, 651) | 2.33 (1.58, 3.42) |
| Jul - Sep | 423 (335, 527) | 0.91 (0.67, 1.24) | 234 (166, 321) | 1.03 (0.66, 1.62) |
| Oct - Dec | 394 (304, 503) | 0.85 (0.62, 1.17) | 50 (20, 103) | 0.22 (0.09, 0.49) |

### Recurrent CHIKF episodes

We determined whether recurrent CHIKF episodes occurred in our study population. To do this we identified all 443 children with first or only episodes of CHIKF and screened all subsequent samples collected during febrile episodes during the period 2014-2018. We defined recurrent episodes as CHIKV RT-PCR positivity during these subsequent febrile dispensary visits. Of the 443 index cases, 170 presented to the dispensary with fever on at least one other occasion during the study period, contributing a total of 320 subsequent febrile dispensary visits with varying durations since the corresponding index CHIKF case (Figure 1). Of the 170 children, 19 (11.2%, 95% CI 6.86, 16.90) were CHIKV RT-PCR positive on at least one subsequent febrile episode (Figures 1 and 2). The duration between the index and the subsequent RT-PCR positive febrile episodes ranged from 2 to 43 months (Figure 2, and Supplementary Material Table S2). Eighteen of the 19 CHIKV RT-PCR positive cases were detected during or after the 2016 CHIKF epidemic year. Neither age, sex nor geographic location were statistically associated with recurrent episodes of CHIKF (Chi2 test p>0.05 for all) and the RT-PCR Ct values and clinical diagnoses were comparable between the index case and corresponding subsequent timepoints (Supplementary Material Figure S2 and Table S2).

**Figure 2:**
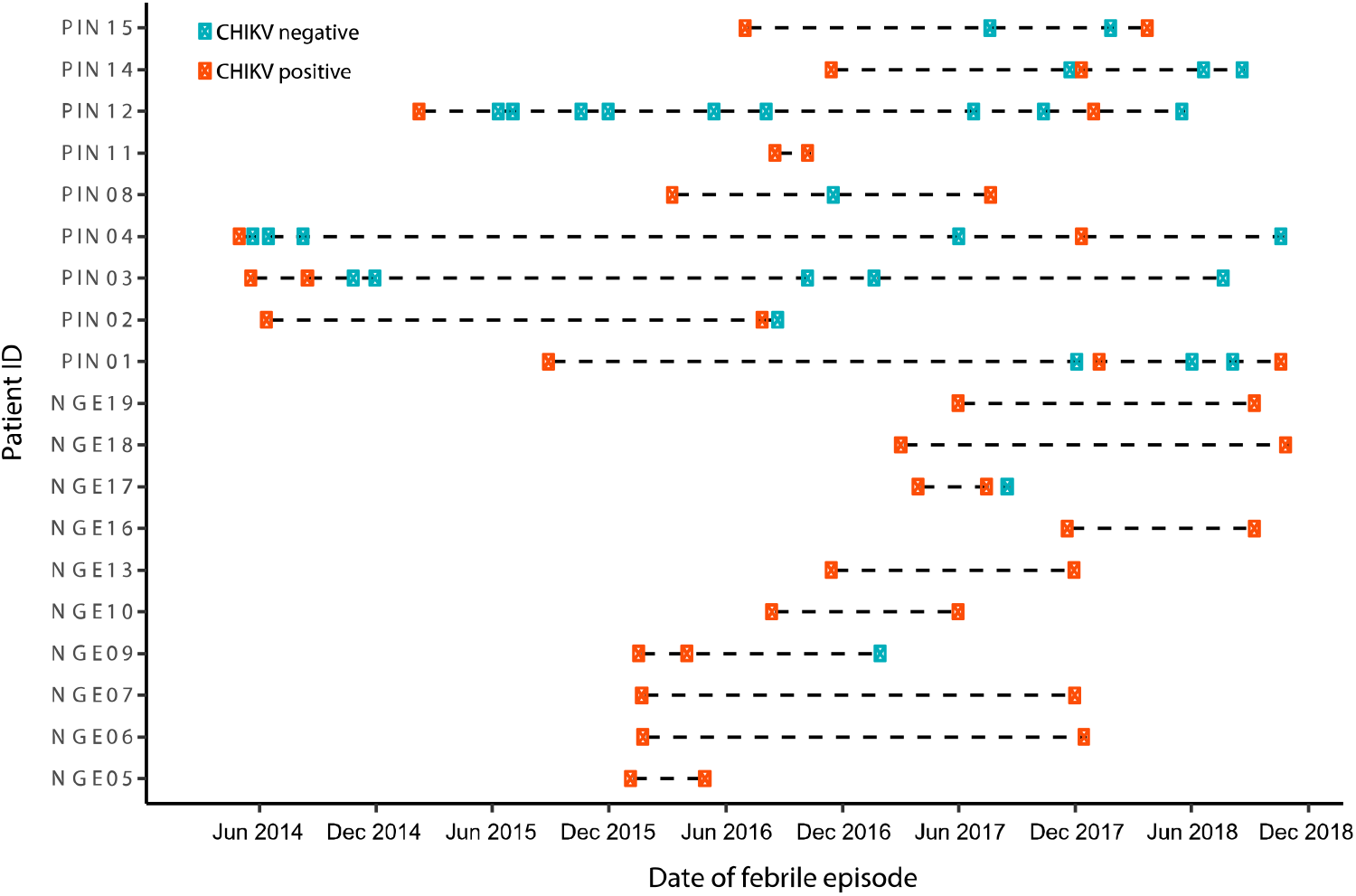
Recurrent CHIKF episodes. The distribution of febrile episodes for each of 19 children who had at least two CHIKV RT-PCR positive febrile episodes is shown. The timing of the CHIKV RT-PCR positive results is shown, with the index episode shown as the first data point in the follow up timeline. Children with prefix ‘NGE’ were resident in Ngerenya and those with ‘PIN’ from Pingilikani, respectively. Further details of these children can be found in Supplementary Material Table S2.

### Viral phylogeny

We attempted to sequence all index and subsequent RT-PCR positive samples from the 19 children with recurrent CHIKF episodes (Figure 2). Complete and partial genome coding sequences were generated from 10 samples, including an additional index sample with a low Ct value (Ct=19) from a child who did not experience recurrent CHIKF episodes during the study period. All CHIKV genomes generated in this study mapped to the ECSA genotype, albeit distinct from the clade containing the IOL strains that emerged during the 2004 epidemic and the clade including genomes from CHIKF patients sampled in Mandera, northern Kenya, during the 2016 epidemic (Figure 3). The E1 A226V mutation, associated with adaptation and increased transmissibility by *Aedes albopictus* [15], was absent from all sequences sampled in Kenya [23]. Time-resolved phylogenies showed strong temporal clustering of CHIKV sequences from coastal Kenya that may suggest antigenic changes over time driven by acquisition of herd immunity (Figure 3). Index-recurrent episode case genome sequence pairs were available for four children with respective time intervals of 2.9, 3.5, 22.3 and 28.3 months between the index and recurrent episodes (Figure 3). The degree of relatedness of the sequence pairs was similarly time-dependent, with the sequence pair with the longest time interval (28.3 months) being most divergent (Figure 3).

**Figure 3:**
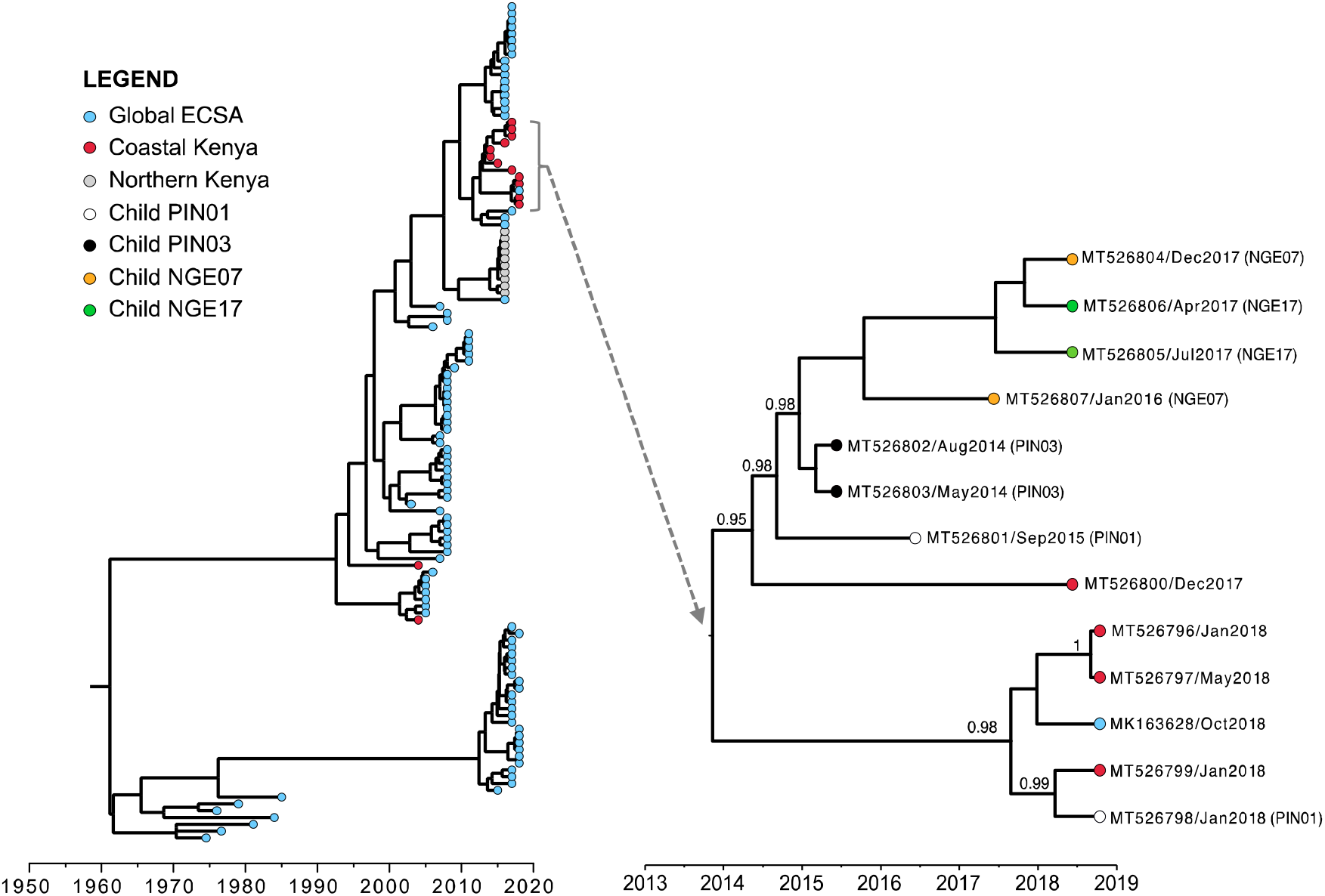
Phylogenetic analysis of CHIKV genomes from Kenya. BEAST MCC tree of 123 ECSA genomes collected across the world, including 24 from Kenya is shown in the left panel. On the right panel is an extraction highlighting the CHIKV sequences generated in this study. The tips are coloured according to geographic location or child identity (as in Figure 2, for those with paired index-recurrent episode sequence pairs), with posterior probability support shown for select nodes with probability >0.8. The same and more detailed MCC tree can be found in the Supplementary Material Figure S1.

## DISCUSSION

In summary, we find that CHIKF is endemic in coastal Kenya with the case burden being particularly high in young children aged <1 year. Our population-based cohort approach in primary healthcare facilities could detect an increase in CHIKF cases during the 2016 epidemic, supporting the utility of such a surveillance framework in the early detection of CHIKF epidemics. During the epidemic year an increase in CHIKF incidence was only noted in Ngerenya while that in Pingilikani (located approximately 40km south of Ngerenya) had been declining [25]; in our dataset, clinical malaria accounted for 57% of all fevers in Pingilikani but only 5% in Ngerenya, while URTI accounted for 70% of all fevers in Ngerenya but only 10% in Pingilikani. We observed significant CHIKF incidence in both locations, and an inverse relationship between CHIKF incidence and age was evident at both locations. This suggests that immunity is acquired in older children, and that CHIKF transmission is significant in both locations despite the varying malaria ecology.

Consistent with previous reports of CHIKF in children in East Africa [19, 30], none of the patients in this study had a clinical diagnosis of CHIKF or reported symptoms of joint pathology. Nineteen children were CHIKV RT-PCR positive during subsequent febrile episodes, with viral genome sequences from these recurrent episodes not showing high degrees of relatedness to the index case. Chronic relapsing and remitting CHIKF symptoms have been observed in adult patients during previous epidemics, as an inflammatory post-infectious arthropathy [31-33] with viral RNA detected in the joint tissues in one study [34]. However, we are aware of no reports of recurrent discrete CHIKF episodes associated with reinfection. This may simply be due to the rarity of longitudinal surveillance studies for CHIKF, particularly in children. In our study, the relatedness of virus genomes from first and second infections in the same child was similar to the relatedness of virus genomes from different children, suggesting that these were reinfections rather than relapses.

Neutralising antibodies that develop following CHIKV infection have been shown to correlate with decreased risk of clinical illness in a longitudinal study in the Phillipines [35, 36], though CHIKF cases in that study were all due to the Asian CHIKV genotype. The possibility that the ECSA genotype viruses, which tend to be more diverse [12] and associated with a higher clinical disease burden than Asian genotypes [37], results in a variable immune response in children in East Africa needs to be ruled out. The inverse relationship between age and incidence of CHIKF suggests that there is an acquisition of protective immunity, although the recurrent infections suggest that this immunity is not rapidly acquired in some children.

Only finger-prick whole blood samples for RT-PCR analysis were available for this study, thus precluding any immunological work. Future longitudinal studies with serial blood sampling for immunological assays will help determine the natural course of infection, including duration of viraemia, and identify host and viral factors that may underlie the occurrence of CHIKF relapses in coastal Kenya.

Consistent with other reports of CHIKV strains circulating in East Africa [12, 23], the viral genomes generated in this study all mapped to the ECSA genotype. Further, in contrast to the 2016 CHIKF epidemic viruses from Mandera (northern Kenya) that formed a single well-supported clade, sequences from coastal Kenya were highly diverse and fell within multiple well-supported clades that whose divergence increased with time. Together, these data suggest ongoing endemic CHIKV transmission at the Kenyan coast possibly sustained through frequent emergence of diverse CHIKV strains from local sylvatic transmission cycles [6].

Our data and that from others in the region [30] further reinforce the need for inclusion of CHIKF high on the list of differential diagnoses to consider when faced with a febrile child at primary healthcare facilities in coastal Kenya. Access to rapid diagnostics would be of value in antibiotic stewardship, and control of transmission in the community would substantially reduce the burden on healthcare facilities. Further studies on the associated clinical outcomes, including the incidence of severe (‘atypical’) manifestations of CHIKF, should help determine the wider public health significance of CHIKF in coastal Kenya.

## Data Availability

The replication data and analysis scripts for this manuscript shall be made available at the Harvard Dataverse: (https://dataverse.harvard.edu/dataverse/kwtrp). Some of the clinical dataset contains potentially identifying information on participants and is stored under restricted access. Requests for access to the restricted dataset should be made to the Data Governance Committee of the KEMRI-Wellcome Trust Research Programme.

## ACKNOWLEDGEMENTS

This manuscript was submitted for publication with permission from the Director of the Kenya Medical Research Institute.

